# The Timing of Macronutrient and Major Food Group Intake and Associations with Mortality Among US Adults, 1999-March 2020

**DOI:** 10.1101/2025.05.27.25328418

**Authors:** Yanbo Zhang, Sarah Alver, Zhilei Shan, Yasmin Mossavar-Rahmani, Ju Zhang, Marie-Pierre St-Onge, Robert Kaplan, Xiaonan Xue, Qibin Qi

## Abstract

**Importance:** Eating timing has been increasingly linked to human health, yet national trends in macronutrient/food group timing and their health implications remain unclear.

**Objective:** To characterize trends in timing of energy, macronutrient, and food group intake among US adults and examine their associations with mortality.

**Design:** Cross-sectional analysis of eating timing trends using National Health and Nutrition Examination Survey (NHANES, 1999-March 2020) data; longitudinal analysis of mortality through December 2019.

**Setting:** A nationally representative sample.

**Participants:** Adults aged ≥20 years with valid 24-hour dietary recall data.

**Exposures:** Timing of energy, macronutrient, and food group intake across predefined 4-hour blocks: 2:00-5:59 am (predawn), 6:00-9:59 am (morning), 10:00 am-1:59 pm (noon), 2:00-5:59 pm (afternoon), 6:00-9:59 pm (evening), and 10:00 pm-1:59 am (midnight).

**Main Outcomes and Measures:** Secular trends in eating timing and mortality.

**Results:** Among 50,264 adults (mean age, 47.5 years; 51.2% women), evening accounted for the largest proportion of daily energy intake (weighted mean proportions across years, 31.9%-33.3%), followed by noon (24.7%-26.8%), afternoon (19.9%-21.8%), morning (13.5%-14.9%), and overnight (midnight and predawn; 5.6%-6.5%), with 23.4%-28.0% of adults consuming foods at midnight; similar distribution patterns were observed for macronutrient and food intake, except whole grain intake peaked in the morning and fruit, egg, and dairy intake distributed more evenly. Over time, energy intake proportions declined at noon and midnight but increased in the afternoon; while the secular trends varied by macronutrients and food groups. On average, fasting started at 8:34-8:51 pm and ended at 8:41-8:52 am. Mean midpoint and duration for energy intake were 2:38-2:48 pm and 11.9-12.2 hours, respectively. Male, non-Hispanic Black, and socioeconomically disadvantaged groups had larger midnight intake proportions and later intake midpoints. Reallocating 5% of daily energy intake from other blocks to midnight was associated with higher cardiovascular mortality (HR, 1.09; 95% CI, 1.02-1.17), driven by carbohydrates and related foods; reallocating 5% to predawn was associated with higher cancer mortality (1.22; 1.05-1.41), driven by proteins and related foods. Each 1-hour delay in the start/end of fasting and intake midpoint was associated with an 8%-9% higher risk of cardiovascular mortality.

**Conclusion and Relevance:** From 1999 to March 2020, US adults consistently consumed the highest energy, macronutrients, and most foods in the evening and started fasting relatively late, with a quarter of adults having midnight consumption. Higher overnight intake and delayed eating timing were associated with higher mortality, particularly for specific macronutrients and foods, highlighting the need to devise optimal eating timing recommendations incorporating food compositions.

**Key Points:** *Question:* What are the eating timing patterns for macronutrients and major food groups among US adults from 1999 to March 2020, and how are they associated with mortality?

*Findings:* In this nationally representative study of 50,264 adults, evening (6-10 pm) consistently contributed to the highest intake proportions of energy, macronutrients, and most food groups, followed by noon (10 am-2 pm), afternoon (2-6 pm), morning (6-10 am), midnight (10 pm-2 am), and predawn (2-6 am). Notably, midnight intake contributed ∼5% of daily energy intake, with a quarter of adults consuming foods at midnight, a nontrivial amount given its potential health risk. The mean durations for energy and macronutrient intake remained ∼12.0 hours, and fasting began after 20:34. Higher midnight and predawn energy intake might be associated with higher mortality; however, associations between eating timing and mortality varied by macronutrients/foods. Males and adults with lower socioeconomic status had higher midnight intake and delayed intake midpoints.

*Meaning:* High evening intake proportions, midnight consumption, and late fasting onset among US adults raise concerns given the health risk of late eating timing, particularly among males, non-Hispanic Black adults, and adults with low socioeconomic status.

## Introduction

Investigating the role of dietary patterns and behaviors for optimal health, i.e., what and when we should eat, is one of the strategic goals outlined in the *2020-2030 Strategic Plan for NIH Nutrition Research*.^1^ As a crucial component of chrononutrition, eating timing is conceptualized by timepoints (when intake occurs), duration (length of intervals between eating episodes), and distribution (proportions of daily intake consumed within specific time blocks).^2^ While emerging evidence links eating timing to cardiometabolic health,^3^ chrononutrition is still in its infancy,^2^ prompting its identification as a critical knowledge gap by the 2025 Dietary Guidelines Advisory Committee^4^ and multiple professional societies.^5,6^

Eating timing remains poorly characterized among US adults, although two studies have described the timing of energy intake based on self-reported eating occasion names (e.g., self-defined breakfast, snack) using National Health and Nutrition Examination Survey (NHANES) data (2009-2014 and 2011-2018).^7,8^ First, beyond energy intake, it is crucial to consider the timing of macronutrient and food group intake, as different dietary components have distinct circadian metabolism patterns^9–11^ and may exert distinct health effects depending on their timing of consumption.^12^ Second, prior studies anchored eating timing to self-reported eating occasions, the definition and timing of which vary substantially across individuals and populations, making it difficult to compare findings across studies and to develop consistent, evidence-based recommendations.^4^ Evidence using clock time as a standardized time anchor remains limited and needed. Third, little is known about the social determinants of eating timing, which are essential for developing targeted interventions to promote health equity.

Moreover, although emerging evidence supports the health benefits of shifting energy intake earlier in the day (e.g., avoiding breakfast skipping and late-night eating) and prolonging fasting intervals,^3^ health effects of macronutrient-/food-group-specific eating timing remain largely unexplored. Therefore, we analyzed NHANES data from 1999 to March 2020 to characterize trends in the timing of energy, macronutrient, and food group intake among US adults based on a comprehensive eating timing framework. We further examined variations in eating timing across sociodemographic groups and explored associations between eating timing and mortality risk.

## Methods

### Study Design and Population

Since 1999, the NHANES has annually employed a stratified, clustered, four-stage, probability sampling design to recruit a representative sample of the noninstitutionalized civilian population living in the 50 states and the District of Columbia.^13^ Data were released in 2-year cycles; however, data collection in 2019-2020 was interrupted by the COVID-19 pandemic, and the 2019-March 2020 and 2017-2018 data were combined to provide nationally representative estimates.^13^

NHANES collected demographic, health, and nutrition information from questionnaire interviews and standardized physical examinations.^13^ The NHANES protocol was approved by the National Center for Health Statistics research ethics review board. All participants provided written informed consent. This analysis included non-pregnant adults aged 20 years and older who completed at least one valid dietary recall (defined in eAppendix in the **Supplement**) across 10 NHANES cycles from 1999-2000 through 2017-March 2020.

### Assessment of Macronutrients and Food Groups

NHANES employed 24-hour dietary recalls using the multiple-pass method and standardized measuring guides to ensure complete and accurate recalls while minimizing respondent burden.^14^ Participants reported the clock time and name (e.g., breakfast, lunch, dinner, snack) of each eating occasion, along with the amount of each food/beverage consumed during that occasion; all foods and beverages consumed on the previous day (midnight to midnight) were thereby documented. All cycles conducted at least one 24-hour dietary recall in person at the Mobile Examination Center; since 2003, a second recall was administered by telephone approximately 3 to 10 days after the first recall.^14^ Macronutrient and energy intake were calculated using the biannually-updated USDA Food and Nutrient Database for Dietary Studies (FNDDS).^15^ Food group servings were estimated using the biannually-updated USDA FPED or MyPyramid Equivalents Database (MPED).^15^

### Eating Timing

Eating timing was conceptualized separately for energy, macronutrients (including carbohydrate, protein, and fat), and major food groups (including whole grains, refined grains, whole fruits, fruit juice, vegetables, red/processed meat, poultry, seafood, eggs, dairy, and lentils/nuts/soy).

Based on a previous study,^16^ the 24-hour cycle was divided into six predefined 4-hour blocks – 2:00-5:59 am (predawn), 6:00-9:59 am (morning), 10:00 am-1:59 pm (noon), 2:00-5:59 pm (afternoon), 6:00-9:59 pm (evening), and 10:00 pm-1:59 am (midnight). To support this segmentation scheme, we calculated the mean hourly intake of energy, macronutrients, and food groups across the day. To further examine potential heterogeneity within each block, we divided each 4-hour block into two equal 2-hour blocks. Self-reported eating occasion names (breakfast, lunch, dinner, and snack) were considered in secondary analyses.

Eating timing has three conceptualizations according to a methodological review (eFigure 1 in the **Supplement**).^2^ First, timepoints described when consumption occurred, including (1) whether each macronutrient/food group was consumed within each block, (2) the clock time of the daily intake midpoint for each macronutrient/food group, and (3) the clock times when fasting for macronutrient intake started and ended (fasting was defined as the longest interval between two eating occasions on survey day). Second, for each macronutrient/food group, distribution was assessed by the proportion of daily intake consumed within each block. Third, durations of energy and macronutrient intake were calculated as the time difference between the end and start of fasting. We did not calculate fasting start/end time or duration for food group intake, as many participants consumed certain food groups once or fewer per day.

**Figure 1.**
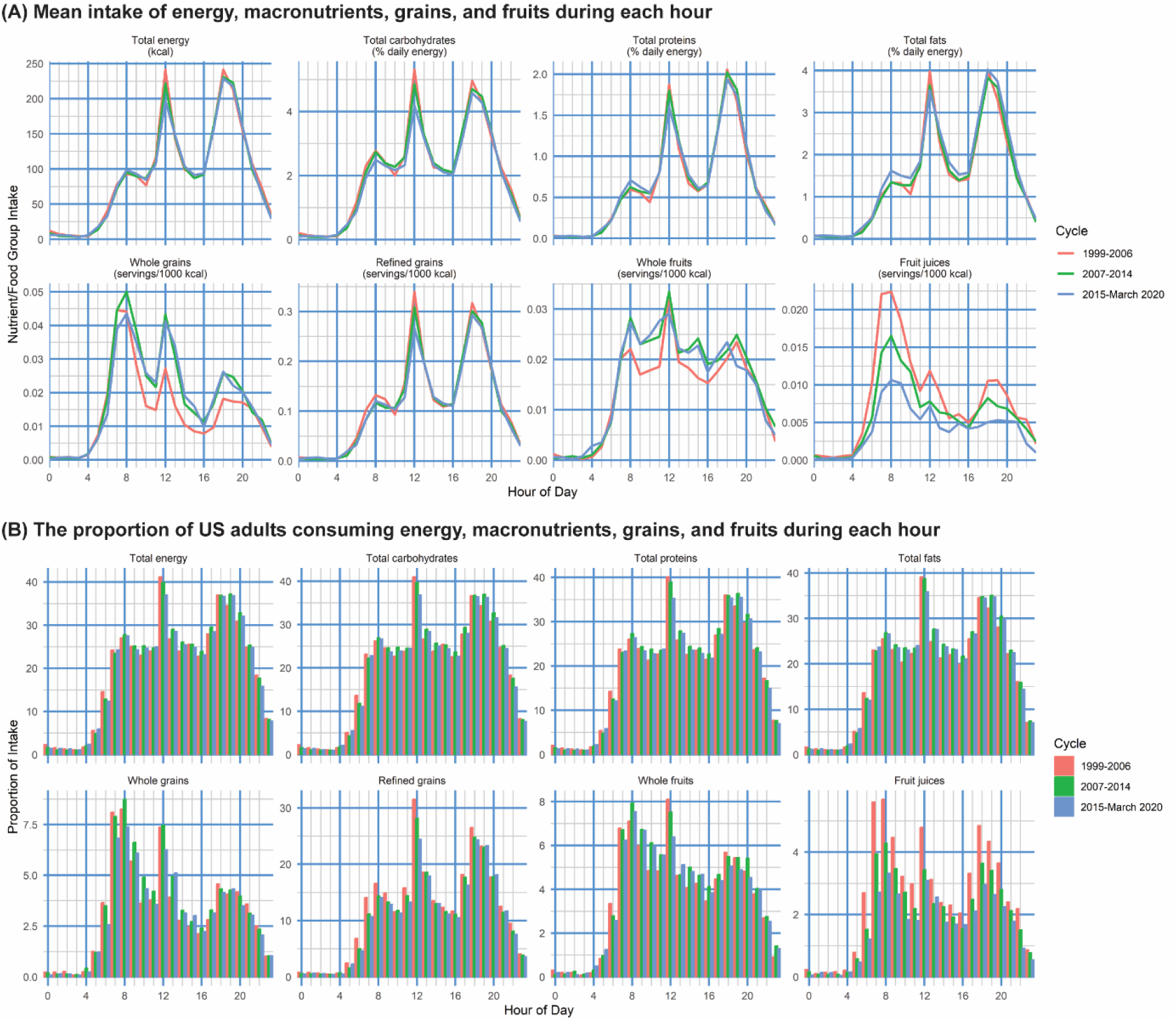
Intake of Energy, Macronutrients, Grains, and Fruits during Each Hour among US Adults, 1999-March 2020. X-axes indicate the hour of the day (in 24-hour format), e.g., 0 indicates 0:00-0:59 am, 16 indicates 4:00-4:59 pm. Y-axes indicate (A) the mean intake of energy, macronutrients, grains, and fruits during each hour or (B) the proportion of US adults consuming energy, macronutrients, grains, and fruits during each hour. Survey years were divided into three groups (1999-2006, 2007-2014, and 2015-March 2020) with roughly similar time spans. Sampling weights, stratification, and clustering of the complex sampling design were considered.

We only used the first dietary recall to estimate population-level eating timing on a given day, under the assumption that the data are accurate when collected evenly throughout the year and the days of the week.^17^ We also performed a sensitivity analysis by averaging two 24-hour dietary recalls among those who completed two recalls.

Several secondary outcomes were considered, including subtypes of carbohydrates (high- and low-quality carbohydrates, defined by food sources; eTable 1 in the **Supplement**), proteins (animal and plant proteins), and fats (saturated [SFAs], monounsaturated [MUFAs], and polyunsaturated fats [PUFAs]),^18^ and mean Healthy Eating Index (HEI) 2020 score within each block (eTable 2 in the **Supplement**).^19^

### Mortality Assessment

Mortality status, date of death, and death cause were obtained via the National Death Index through December 31, 2019. We examined mortality from all causes, cancer (International Classification of Diseases, 10th Revision codes, C00-C97), and cardiovascular diseases (CVDs; I00-I09, I11, I13, I20-I51, and I60-I69). To reduce the potential for reverse causation, participants with fewer than five years of follow-up were excluded.^20^

### Statistical Analysis

All analyses considered sampling weights, stratification, and clustering of the complex sampling design to derive nationally representative estimates. The sampling weights considered race and Hispanic origin, age group, sex, and weekday/weekend for dietary recalls. First, for each macronutrient and food group, each eating timing conceptualization was described by survey cycles, and weighted means or proportions with 95% confidence intervals (CIs) were estimated. To illustrate the secular trend of eating timing, we constructed two general linear regression models by regressing each eating timing variable on the survey cycle (detailed in eAppendix in the **Supplement**); survey cycle was treated as a continuous variable (to estimate *P* values for overall secular trends) and a categorical variable (to calculate absolute differences between the 2017-March 2020 and 1999-2000 cycles), respectively.

Second, we explored social determinants of eating timing by describing the timing of macronutrient and food group intake in different subgroups (by age, sex, race/ethnicity, education, income, employment status, and food security; detailed in eAppendix in the **Supplement**).

Third, Cox regression models estimated hazard ratios (HRs) and 95% CIs for mortality associated with eating timing, i.e., for each macronutrient/food group, consumption within each block (yes vs no), daily intake midpoint (per hour delay), fasting start/end time (per hour delay), proportion of daily intake within each block (per 5% increase), and duration (per hour increase).

Models adjusted for age, sex, race/ethnicity, education, income, employment status, food security, daily energy intake, daily intake of the examined macronutrient/food group, number of daily eating occasions, daily HEI score, sleep duration, cigarette smoking, alcohol consumption, leisure-time physical activity, body mass index, and prevalent hypertension, high cholesterol levels, diabetes, cardiovascular disease, and cancer. Carbohydrate, protein, and fat intake within each block were mutually adjusted due to high correlations (r range, 0.48-0.83). Within each block, food group intake with high correlation was also mutually adjusted (eAppendix in the **Supplement**). Unlike other analyses examining population-level distributions, this analysis examined individual-level eating timing. Thus, to improve the reliability of eating timing estimates for each participant, only participants with two 24-dietary recalls were included, with data averaged across two days;^21^ two-day dietary sampling weights were thereby used to generate nationally representative estimates.^13^ All data were analyzed using R version 4.4.1 (The R Foundation). Two-sided *P* values of <.05 were considered statistically significant.

## Results

### Study Design and Population

Between 1999 and March 2020, 51,688 NHANES participants aged 20 years or older provided at least one valid dietary recall. After excluding 1,424 pregnant participants, 50,264 participants (weighted mean age, 47.5 years [SE, 0.19 years]; 25,326 [51.2%] females) were included in this analysis. From 1999 to March 2020, the mean age increased from 46.1 to 48.7 years (eTable 3 in the **Supplement**); the proportion of non-Hispanic White adults declined from 71.4% to 64.2%; there were increasing trends in socioeconomic levels, but the prevalence of low/very low food security increased.

### Timing of Energy Intake

Between 1999 and March 2020, three energy intake peaks were consistently observed at 8:00- 8:59 am, 12:00-12:59 pm, and 6:00-6:59 pm, with similar peaks for macronutrients and most food groups (**Figure 1** and eFigure 2 in the **Supplement**). The first two peaks occurred near the midpoint of predefined morning (6:00-9:59 am) and noon (10:00 am-1:59 pm), while the third aligned with the onset of evening (6:00-9:59 pm). Furthermore, across twelve 2-hour blocks, four blocks spanning 10:00 pm-5:59 am contributed the least to macronutrient and food group intake (eFigures 3-4 in the **Supplement**), corresponding to two blocks with minimal intake: midnight and predawn (collectively referred to as overnight). These findings support the rationale for the current 4-hour block structure.

**Figure 2.**
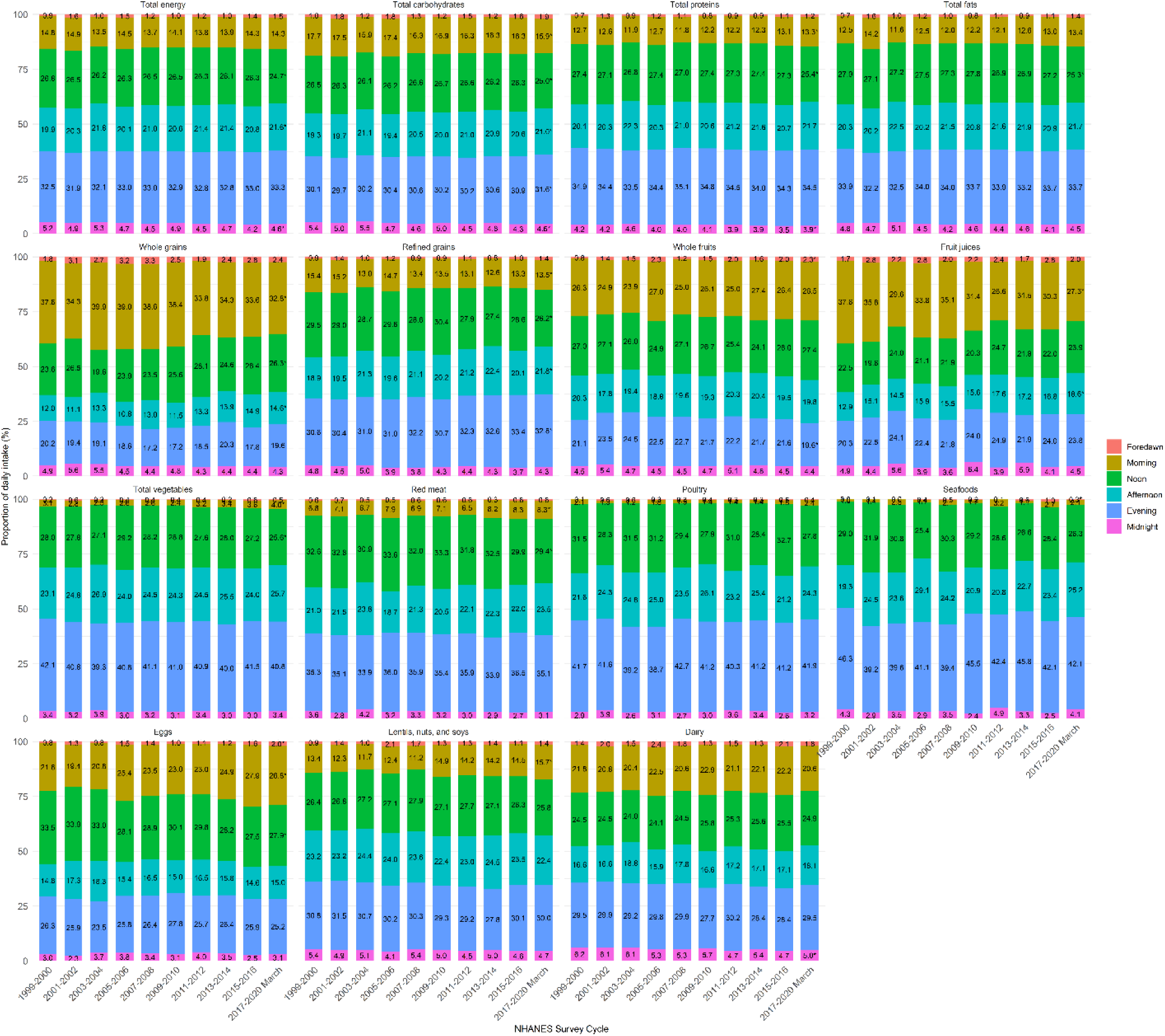
Proportion of Macronutrient and Food Group Intake in Each Block among US Adults by National Health and Nutrition Examination Survey Cycles from 1999 to March 2020. Sampling weights, stratification, and clustering of the complex sampling design were considered. For each nutrient or food group, the intake proportion at each block was calculated among participants who reported consuming the certain nutrient or food group on the survey day. Six 4-hour blocks were predefined – 2:00-5:59 am (predawn), 6:00-9:59 am (morning), 10:00 am-1:59 pm (noon), 2:00-5:59 pm (afternoon), 6:00-9:59 pm (evening), and 10:00 pm-1:59 am (midnight). * *P* for trend <.05. The difference between 2017-March 2020 and 1999-2000 and *P* for trend are reported in eTable 4 in the **Supplement**.

**Figure 3.**
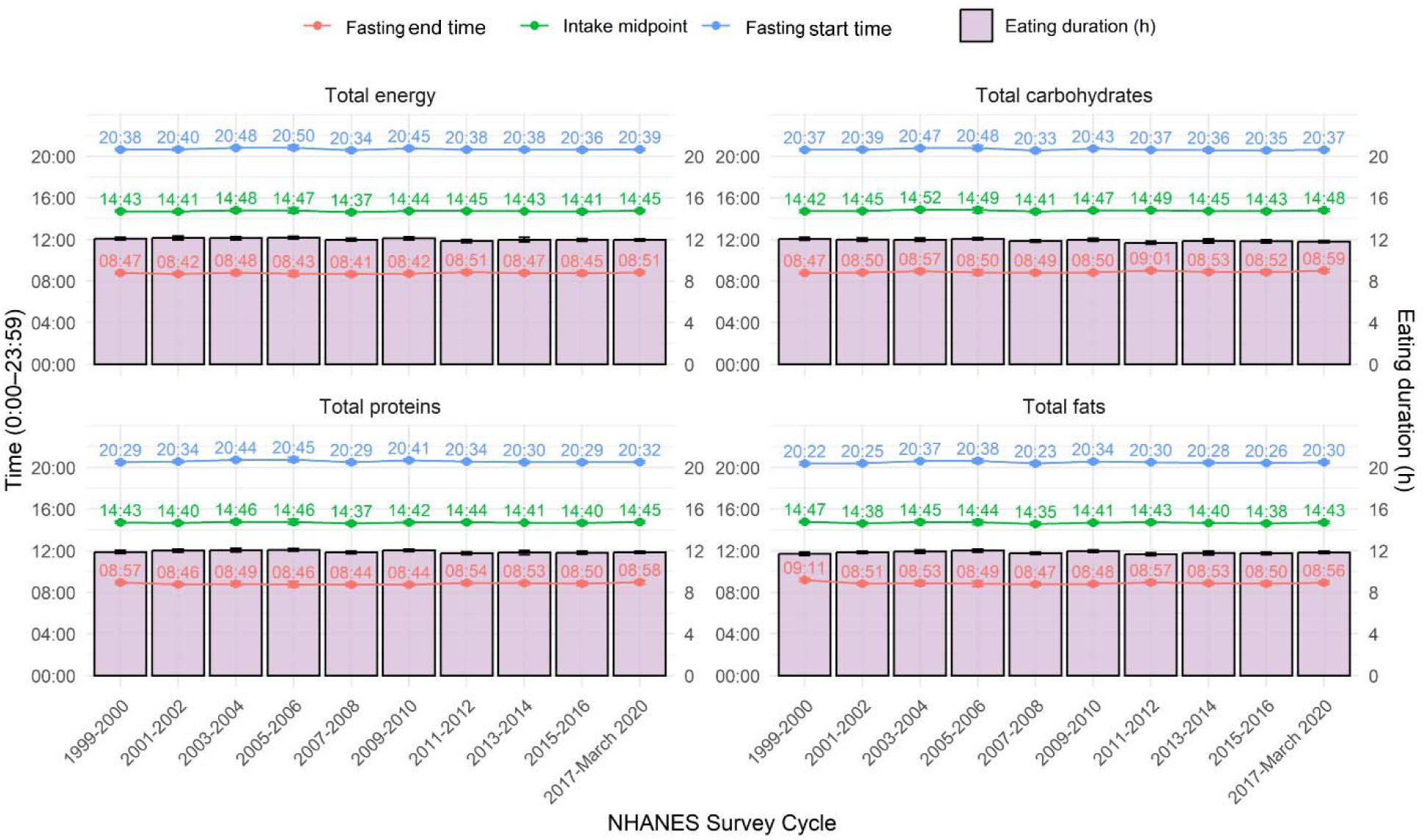
Fasting End and Start Time, Intake Midpoints, and Eating Durations for Energy and Macronutrients among US Adults by National Health and Nutrition Examination Survey (NHANES) Cycles from 1999 to March 2020. X-axes denote the NHANES survey cycle; left y-axes denote the clock time (in 24-hour format, e.g., 16:00 means 4:00 pm) for the mean fasting end/start time and intake midpoints, corresponding to the dots in the figure; right y-axes denote the eating duration (hours), corresponding to the bar in the figure. Sampling weights, stratification, and clustering of the complex sampling design were considered. Only participants who consumed certain macronutrients on the survey day were included. Between March 2020 and 1999, difference in fasting end time for carbohydrate intake was 0.20 (95% confidence interval, 0.01, 0.39) hours; difference in carbohydrate intake duration was -0.26 (-0.42, -0.10). There are declining trends in energy and protein intake duration (*P*<.003 for trend), and the difference between March 2020 and 1999 was -0.12 (-0.28, 0.04) hours and -0.04 (-0.22, 0.13) hours, respectively. No secular trends were observed for other parameters.

**Figure 4.**
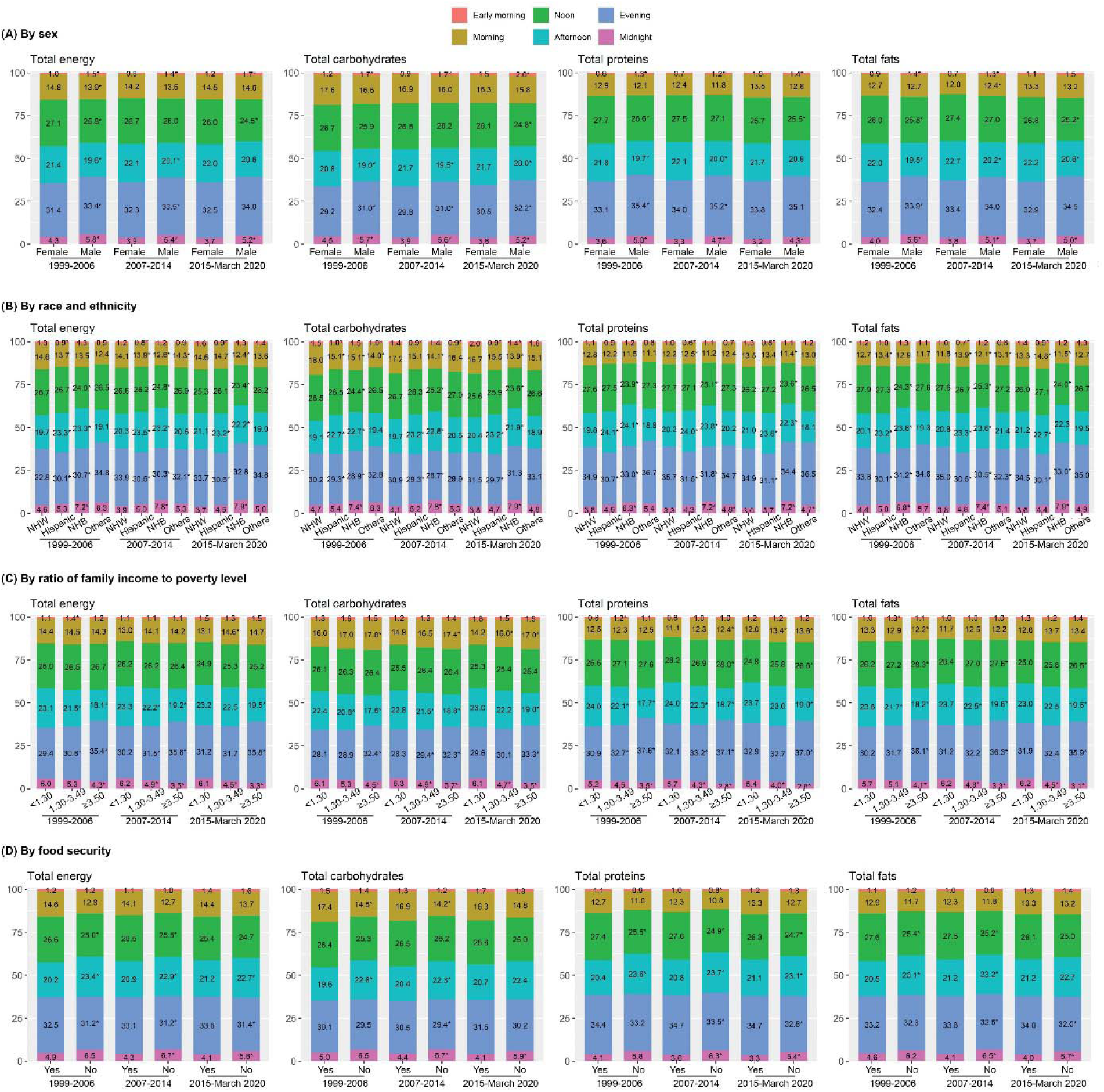
Proportion of Energy and Macronutrient Intake at Each Block by Sex, Race/Ethnicity. Income, and Employment Status among US Adults, 1999–March 2020. Sampling weights, stratification, and clustering of the complex sampling design were considered. Six 4-hour blocks were predefined – 02:00-05:59 (predawn), 06:00-09:59 (morning), 10:00-13:59 (noon), 14:00-17:59 (afternoon), 18:00-21:59 (evening), and 22:00-01:59 (midnight). Results for food intake and other subgroups are shown in **Supplement**. * *P* for interaction < .05.

From 1999 to March 2020, over 82.5% of US adults consumed energy in the evening and noon, respectively; over 70.5% in the morning and afternoon, respectively; 23.4%-28.0% at midnight; 7.6%-10.7% in the predawn (eFigure 5 in the **Supplement**). US adults consumed the highest proportion of daily energy in the evening (mean proportions across 1999-March 2020, 31.9%-33.3%), followed by noon (24.7%-26.8%), afternoon (19.9%-21.8%), morning (13.5%-14.9%), midnight (4.2%-5.3%), and predawn (0.9%-1.6%; **Figure 2**).

**Figure 5.**
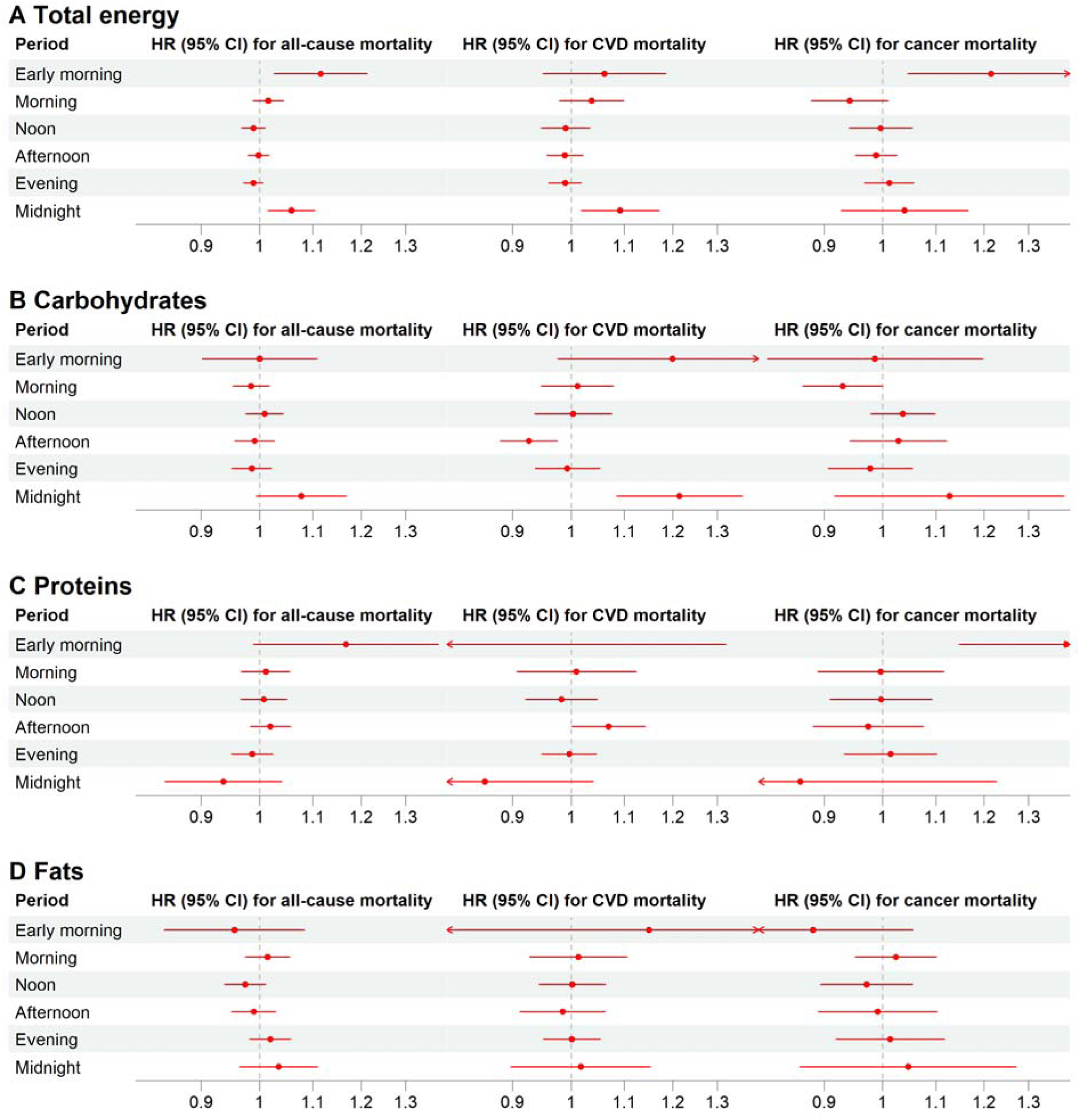
Associations of Distribution for Energy and Macronutrient with Mortality. Sampling weights, stratification, and clustering of the complex sampling design were considered. Analyses only included participants recruited in 2005-2014, because 1) participants with less than 5-year follow-up were excluded from the analyses to reduce the possibility of reverse causation, and those recruited after 2014 were excluded (mortality data were updated until the end of 2019); 2) only participants providing two 24-hour dietary recalls were included to more accurately reflect participants’ dietary habits, and the National Health and Nutrition Examination Survey (NHANES) only administered one 24-hour dietary recall for each participant before 2003; 3) the NHANES started to collect sleep information in 2005. Red dots with horizontal lines indicate the hazard ratios (HRs) with their 95% confidence intervals (CIs) related to per 5% increase in the intake in each block to daily intake. We adjusted for age, sex, race/ethnicity, education, income, employment status, food security, daily energy intake, daily intake of the examined macronutrient, number of eating occasions, Healthy Eating Index-2020 score, sleep duration, cigarette smoking, alcohol consumption, leisure-time physical activity, body mass index, and prevalent hypertension, high cholesterol levels, diabetes, cardiovascular disease, and cancer. Macronutrient intake at each block was mutually adjusted given high correlations (r range, 0.48-0.83).

Over these years, mean proportion of daily energy intake consumed at noon (from 26.8% to 24.7%) and midnight (from 5.2% to 4.6%) decreased, while increasing from 19.9% to 21.6% in the afternoon (*P*≤.028 for trend; **Figure 2** and eTable 4 in the **Supplement**), with no statistical change at other blocks. Same secular trends were observed for the proportion of adults with energy intake within each block (eFigure 5 in the **Supplement**).

Consistently, self-reported dinner contributed to the highest proportion of energy intake (35.6%-36.6% across years), followed by lunch (24.3%-25.8%), snack (20.9%-23.6%), and breakfast (15.3%-17.3%; eFigures 6-7 in the **Supplement**). Results were largely consistent when using the average from two dietary recall data (eFigure 8 in the **Supplement**).

From 1999 to March 2020, fasting consistently ended at 8:41-8:52 am and started at 8:34-8:51 pm. The midpoint of energy intake remained stable at 2:38-2:48 pm; energy intake duration ranged 11.9-12.2 hours (**Figure 3**).

### Timing of Macronutrient Intake

Within each block, the proportion of adults reporting intake of each macronutrient was similar to that of energy intake (eFigure 5 in **Supplement**). Similar to energy intake, US adults consumed the highest proportion of daily macronutrient intake in the evening, followed by noon, afternoon, morning, midnight, and predawn between 1999 and March 2020 (**Figure 2**). For example, in 2017-March 2020, 31.6% of daily carbohydrates occurred in the evening, 25.0% at noon, 21.0% in the afternoon, 15.9% in the morning, 4.6% at midnight, and 1.9% in the predawn; within each block, proportions of carbohydrates, protein, and fat consumed relative to their respective daily intake totals were similar.

However, secular trends in eating timing varied by macronutrients. From 1999 to March 2020, the proportion of daily carbohydrate intake slightly decreased in the morning, noon, and midnight, while increasing in the afternoon and evening; all changes were within ±1.8% (*P*≤.021 for trend; **Figure 2** and eTable 4 in the **Supplement**). A similar trend was observed for low-quality rather than high-quality carbohydrates (eFigure 9 in the **Supplement**). In contrast, there were declining trends in proportion of daily protein (from 27.4% to 25.4%) and fat (from 27.9% to 25.3%) intake at noon, with minimal changes in proportions at other blocks (**Figure 2**). For protein subtypes, the proportion of daily animal protein intake at noon decreased over these years, while the noon proportion of plant protein intake remained stable; additionally, the morning proportion of animal protein intake increased, while that of plant protein intake decreased (eFigure 9 in the **Supplement**). For all fat subtypes, the proportions of daily intake increased in the morning but decreased at noon over these years. Within each block, carbohydrates consistently contributed to the highest energy intake, followed by fats and proteins (eFigure 10 in the **Supplement**).

**Figure 3** shows that fasting start/end time, intake midpoint, and eating duration for macronutrients are similar to those of energy intake and largely stable between 1999 and March 2020. However, the mean fasting end time for carbohydrate intake was delayed by 12 minutes (from 8:48 to 9:00 am over these years; *P*=.045), which shortened the mean carbohydrate intake duration by 0.3 hours (from 12.1 hours to 11.8 hours; *P*=.002).

### Eating Timing for Major Food Groups

For refined grains, total vegetables, red/processed meat, poultry, seafood, and lentils/nuts/soy, US adults consumed the highest proportion in the evening, followed by noon, afternoon, morning or midnight, and predawn (**Figure 2**). Particularly, morning, midnight, and predawn collectively contributed to <12% of daily intake for total vegetables, red/processed meat, poultry, and seafood, and <12.9% adults consumed these food groups during these three blocks (eFigure 5 in the **Supplement**). In contrast, US adults consumed the largest proportion of whole grains in the morning (range, 32.8%-39.9%), followed by noon (19.6%-28.1%), evening (17.2%-20.3%), afternoon (10.8%-14.9%), midnight (4.3%-5.6%), and predawn (1.8%-3.3%). Whole fruit, fruit juice, egg, and dairy intake showed more balanced distributions across morning, noon, and evening, e.g., 23.9%-27.4%, 24.1%-27.4%, and 19.6%-24.5% for whole fruit intake, respectively. The mean HEI score ranged 29.6-39.4 across morning, noon, afternoon, and evening and 3.0-11.2 across predawn and midnight (collectively referred to as overnight; eFigure 11 in the **Supplement**), suggesting low dietary quality during the overnight period.

### Social Determinants of Eating Timing

Eating timing distribution for macronutrients and major food groups is largely consistent across age groups (eFigure 12 in the **Supplement**); however, adults aged 35-64 years had longer durations for macronutrient intake and ended fasting earlier compared to those aged 20-34 years (eFigure 13 in the **Supplement**). Compared to females, males consumed greater proportions of energy and macronutrients in the evening, midnight, and predawn, but smaller proportions at noon and afternoon (**Figure 4** and eFigure 14 in the **Supplement**); males had an overall later eating window (later fasting end/start time and intake midpoint) and a longer macronutrient intake duration (eFigure 15 in the **Supplement**).

Compared to non-Hispanic White adults, Hispanic adults consumed larger proportions of energy and macronutrients in the afternoon, larger proportions of fats in the morning, and smaller proportions of energy and macronutrients in the evening and predawn (**Figure 4** and eFigure 16 in the **Supplement**); they ended fasting later and thus had shorter durations for all macronutrient intake (eFigure 17 in the **Supplement**). Non-Hispanic Black adults consumed smaller proportions of energy and macronutrients in the morning, noon, and evening, while larger proportions in the afternoon and midnight; they had overall later eating windows (later fasting end/start time and intake midpoint) and shorter eating durations for all macronutrients.

Generally, adults with favorable socioeconomic factors (i.e., high income, employment, and food security) ended their fasting earlier but had similar fasting start times, compared to their counterparts (eFigures 18-21 in the **Supplement**); consequently, they had earlier intake midpoints and longer durations for all macronutrients. They also consumed greater proportions of energy, macronutrients, and most food groups in the evening but smaller proportions in the afternoon or midnight (**Figure 4** and eFigures 22-25 in the **Supplement**), and they consumed greater proportions of whole fruits, lentils/nuts/soy, or whole grains in the morning.

### Associations between Eating Timing and Mortality

Among 20,730 NHANES participants with two 24-hour dietary recalls (2005-2014), 1,783 deaths (520 from CVD, 409 cancer) were documented over 200,802 person-years (median=9.7 years).

Several findings supported higher mortality risk associated with delayed eating timing (**Figure 5** and eFigure 26 in the **Supplement**). First, reallocating 5% of daily energy intake from other blocks to midnight or predawn was associated with higher mortality. HRs (95% CIs) per 5% increase at midnight were 1.06 (1.01-1.10) for all-cause, 1.09 (1.02-1.17) for CVD, and 1.04 (0.93-1.17) for cancer mortality; the corresponding HRs for predawn energy intake were 1.12 (1.03-1.21), 1.06 (0.95-1.19), and 1.22 (1.05-1.41), respectively. Particularly, higher midnight intake proportions for carbohydrates, refined grains, whole fruits, and fruit juices were associated with higher CVD mortality; higher predawn intake proportions for proteins, red/processed meat, poultry, eggs, refined grains, vegetables, and fruit juices were associated with higher cancer mortality. These results were largely consistent with comparisons between consumers and non-consumers at midnight and predawn.

Second, for energy intake, delayed fasting end and start times and intake midpoint were associated with higher CVD mortality, with HRs (95% CIs) per 1-hour delay of 1.08 (1.02, 1.15), 1.09 (1.02, 1.16), and 1.09 (1.02, 1.16), respectively (eFigure 26 in the **Supplement**); similar associations were observed for all-cause mortality. Particularly, higher CVD mortality was only linked to delayed fasting start time and intake midpoint for proteins, but not for carbohydrates or fats. Third, consuming proteins, poultry, eggs, and refined grains in the morning was associated with lower cancer mortality.

Notably, associations between afternoon intake proportions and CVD mortality varied by macronutrient. Reallocating 5% of daily intake from other blocks to afternoon was associated with an HR (95% CI) of 0.93 (0.88, 0.98) for carbohydrates while 1.07 (1.00, 1.14) for proteins. Results remained similar after excluding participants with evening/night/rotating shifts (data not shown).

## Discussion

Over the past two and half decades, US adults consistently consumed the largest proportions of energy, macronutrients, and most foods in the evening, followed by noon, afternoon, morning, midnight, and predawn. Mean durations for energy and macronutrient intake were ∼12.0 hours. Previous studies using NHANES data from 2009-2014^8^ and 2011-2018^7^ reported similar patterns, including the highest energy intake at dinner and a mean eating duration exceeding 12 hours.

However, by dividing the 24-hour cycle into six 4-hour blocks based on clock time, our study overcomes the limitation of self-reported eating occasions that hinders comparability across studies^4^. Supported by prior research^16^ and energy intake peaks observed in this study, this method established a standardized time classification to enhance comparability across future chrononutrition studies. Besides, our study extends these findings by analyzing a longer and more recent timeframe (1999-March 2020) and employing a comprehensive framework that examines the eating timepoint, timing distribution, and duration.

Our findings raised concerns about eating timing patterns among US adults. Chrononutrition research increasingly highlights cardiometabolic risk of late-night eating and lower energy intake in the morning.^3^ Consistently, we observed that higher overnight intake of energy and several macronutrients/foods was associated with higher mortality risk, and consuming proteins, poultry, eggs, and refined grains in the morning was associated with lower cancer mortality. Although midnight energy intake showed a decreasing trend, it still contributed ∼5% of daily intake, with 23.4% and 10.7% of adults consuming foods at midnight and predawn, and the fasting occurred after 20:34, potentially raising concerns about late-night eating.

Moreover, we found that the evening consistently accounted for the highest proportion of daily energy, macronutrient, and most food group intake, with an increasing trend in the proportion of evening carbohydrate intake. On the other hand, morning only contributed to <15% of daily energy intake.

By investigating the timing of macronutrient and food group intake, our study offered additional insights beyond those provided by prior studies that focused on energy intake timing, highlighting two reasons why timing of specific dietary components matters. First, certain food groups have unique eating timing patterns, e.g., disproportionately higher proportions of whole grain and whole fruit intake occurred in the morning among US adults. These findings prompt a reconsideration of previously reported cardiovascular benefits associated with greater morning energy intake,^3^ as it remains unclear whether such benefits are attributable to greater consumption of healthy foods in the morning (compared to other blocks) or to the timing itself.

In our analysis, which adjusted for daily macronutrient/food intake and intake of macronutrients and correlated foods at each block, we found that only higher morning intake of carbohydrates, refined grains, and eggs was associated with lower cancer mortality, rather than total energy or other nutrients/foods, consistent with previous studies.^12,22^ This suggests that reallocating carbohydrate and egg intake to the morning may be more effective in health improvement than merely increasing the morning energy intake proportion.

Second, the health effects of eating timing may differ by nutrient/food group, even within the same time window. Particularly, our study observed opposite directions for associations of afternoon carbohydrate (protective) and protein (risk) intake with CVD mortality. Individuals with higher afternoon carbohydrate intake tended to consume fewer carbohydrates later in the day (eFigure 27 in the **Supplement**); studies have demonstrated lower insulin sensitivity and higher postprandial glucose levels in the evening,^9^ which may explain the benefits of shifting carbohydrate intake from evening/midnight to afternoon. Meanwhile, amino acid metabolisms are more active in the afternoon,^11^ and branched-chain amino acid metabolites have been linked to cardiometabolic risk,^23^ which may explain the association between afternoon protein intake and CVD mortality.

Previous studies have shown that males, non-Hispanic Black individuals, and socioeconomically disadvantaged adults tend to have less healthy diets defined by dietary quantity and quality,^18,24^ and our findings add new evidence from the eating timing perspective – these populations also tended to consume a greater proportion of their daily intake later in the day and have a later intake midpoint, which are associated with adverse health outcomes.

Particularly, both this and previous studies^7,8^ observed shorter eating durations among Hispanic and non-Hispanic Black adults compared to non-Hispanic White adults, but our study suggested different causes – Hispanic adults ended fasting later but started fasting at a similar time, while non-Hispanic Black adults had both delayed fasting end and start times. Correspondingly, Hispanic adults had smaller intake proportions in the evening and predawn, whereas non-Hispanic Black adults had lower morning and higher midnight intake, which were linked to adverse health outcomes.

This study’s strengths include applying a comprehensive eating timing framework, examining macronutrient- and food-group-specific eating timing, leveraging nationally representative data collected during an extensive period, and considering multiple social features. However, several limitations should be acknowledged. First, although supported by prior research^16^ and observed energy intake peaks, the time classification of six predefined 4-hour blocks needs validation from other studies for future application. Second, although 24-hour dietary recall is the most feasible way to collect eating timing data currently, measurement errors are inevitable, and affordable wearable devices combined with artificial intelligence techniques are warranted in future studies for more accurate dietary evaluation. Third, for the mortality analysis, although evidence reported moderate reliability in estimating the eating timing for each individual by using two dietary recalls, using more days of recall data can improve the reliability.

## Conclusions

From 1999 to March 2020, US adults consumed the largest proportions of energy, macronutrients, and most foods in the evening, followed by noon, afternoon, morning, and overnight. A quarter of adults had midnight consumption, with a relatively late fasting start (after 8:34 pm). Higher overnight consumption might be associated with higher mortality, though associations varied by nutrients and foods, underscoring the need for future chrononutrition studies on specific nutrients and foods to establish evidence-based eating timing recommendations. Adults with lower socioeconomic status had greater midnight intake and delayed eating timing, suggesting nutritional disparities in eating timing.

## Data Availability

Data from the NHANES are available at https://wwwn.cdc.gov/nchs/nhanes/default.aspx.

